# A Survey of the Smokefree Status of Pedestrian-Only Spaces in 10 New Zealand Local Government Areas

**DOI:** 10.1101/2021.06.09.21258659

**Authors:** Nick Wilson, Niveditha Gurram, Leah Grout, George Thomson

**Affiliations:** Department of Public Health, University of Otago Wellington; Te Hiringa Hauora/Health Promotion Agency, Wellington

## Abstract

**Aim:** To describe the smokefree status and signage of outdoor pedestrian-only plazas/malls/boulevards in 10 New Zealand local government (council) areas.

**Methods:** The 10 council areas were a convenience sample. Council websites were examined for smokefree policies and a systematic attempt was made to identify the five largest pedestrian-only sites with permanent seating in each council area (10 sites each for two larger cities). Field visits were conducted to all selected sites.

**Results:** Smokefree policies with components covering smokefree outdoor plazas/malls/boulevards were common (80%; 8/10 councils), albeit with some gaps (eg, around signage and vaping policy). A total of 60 relevant pedestrianised sites with permanent seating were identified and surveyed. Of these 63% were officially designated smokefree. Smokefree signage was only present in 15% (9/60) of all the sites and in 24% (9/38) of the designated smokefree sites. In these designated sites the average number of smokefree signs was only 1.4 (range: 0 to 14). Issues identified with the signs included small size, being only a small part of a larger other sign, limited use of te reo Māori wording, and not covering vaping. At sites where tables were present, 12% had ash trays on the tables (none where smokefree).

**Conclusions:** Smokefree plazas/malls/boulevards in this survey had multiple policy and signage deficiencies that are inconsistent with achieving the national smokefree goal for 2025. There is scope to address these issues with an upgrade to the national smokefree law.

## INTRODUCTION

One mechanism to make progress towards reducing the enormous health burden from tobacco smoking, is to expand smokefree public areas. For many high-income countries, this now means an increased focus on outdoor public settings, where smokefree policies are much less common than for indoor public settings. Such policies are intended to reduce the exposure of workers and the public to tobacco smoke pollution and contribute to the denormalisation of smoking by reducing its visibility.^1 2^ A perception of smokefree parks or outdoor dining policies has been associated with increased quit attempts,^3^ and not being exposed to smoking in Ontario bar/restaurant outdoor areas increased quit attempts and decreased smoking relapses.^4^ In New York, smokefree policies for parks and beaches were followed by reduced tobacco-related litter, observed smoking and public perceptions of smoking.^5 6^ In the Netherlands the introduction of an inner-city outdoor smokefree zone was associated with a substantial decline in the number of smokers in the zone.^7^

There is often majority public support for a number of types of outdoor smokefree areas (eg, based on surveys in the USA and Canada,^8^ Spain,^9^ Australia,^10 11^ and the UK^12^). There is even majority support for some outdoor smokefree areas by smokers (eg, in New Zealand,^13^ Spain,^9^ Italy,^14^ Australia,^11^ and North America^8^).

Internationally, the most progress towards outdoor smokefree policy has generally been made in areas explicitly associated with children (eg, schools, playgrounds, urban parks),^15^ along with some movement with outdoor hospitality areas (eg, outside bars and cafés).^16^ However, smokefree policies for central urban pedestrian areas have increased in California,^17^ and some larger Australian cities.^18^ New York has smokefree policies for ‘Pedestrian plazas such as those at Times Square and Herald Square’.^19^

In New Zealand, exposure to second-hand smoke is a serious problem with an estimated 347 premature deaths per year in 2019.^20^ When morbidity from this smoke is also considered, this health loss amounted to an estimated 9022 lost disability-adjusted life years (DALYs) in 2019.^20^ Fortunately, there has been some progress with voluntary or ‘educational’ outdoor smokefree areas over the last two decades.^21-23^ Nevertheless, various issues have been identified in terms of the extent of coverage and quantity/quality of the signage (eg, hospitality settings,^24^ schools,^25^ childrens’ playgrounds,^26^ hospital grounds,^27^ racecourses and sports facilities,^28^ railway stations,^29^ airports,^30^ and various other settings^31^). These limitations are problematic in the context of the country having a Smokefree Goal for the year 2025.^32^

Despite the research mentioned above, there has been little work in New Zealand on evaluating the extent of smokefree pedestrian-only areas (plazas/malls/boulevards). In Rotorua’s pedestrian “Eat Street”, in 2019 a count of smokers 12 months after the adoption of a smokefree policy found relatively few people smoking (0.6% of those observed; ie, 36/6530; personal communication). The measurement of the prevalence of smoking in a few such places has occurred in Wellington, either before or after the adoption of smokefree policies.^33 34^

Given this background, and the revived New Zealand Government interest in progressing tobacco control,^35 36^ we aimed in this study to describe the smokefree status and signage of outdoor pedestrian-only plazas/malls/boulevards in 10 New Zealand local government (council) areas.

## METHODS

### Sample selection

The 10 council areas were a convenience sample based on author living locations and travel plans (north to south the areas were: Hastings, Napier, Palmerston North, Masterton, South Wairarapa, Porirua, Upper Hutt, Hutt, Wellington in the North Island; and Queenstown-Lakes in the South Island). A systematic attempt was made to identify all of the outdoor pedestrian-only plazas/malls/boulevards with permanent seating in these council areas. We then selected the five largest of these by (measured by paved/gravel area) within each council area (or 10 in the case of councils with 100,000+ populations: Wellington and Hutt Cities).

### Smokefree policies

The website of each council was examined to identify the smokefree policy. Key features of the each policy were documented, particularly how it related to the selected sites.

### Site inclusion criteria

For the purposes of this study we used the following definitions for site inclusion in the survey:

- An outdoor plaza with seating was an outdoor area that was at least 50% paved or gravel surfaced, had some distinguishing structures to prevent vehicle access (eg, bollards or planter boxes) and had at least one permanent seat. In some cases these plazas were still officially called “parks” eg, “Grey Street Pocket Park” in Wellington City. Also, we identified plaza areas within larger park settings, where the plaza was well defined by surrounding structures. Where a plaza or boulevard was an extended area (or series of connected areas) of the footpath, we required that the area including the extension was at least twice the width of the nearest normal width of footpath. We took a broad interpretation of seating to include seating with no back support (eg, concrete block or wooden benches that could be sat on).
- A pedestrianised mall/boulevard was defined as an outdoor area that was fully pedestrianised for its entire width, was wider than a typical footpath and which had at least one permanent seat.

### Site identification

For each city the following steps were performed:

- We examined Google Maps, Google Street View, and undertook our own observations within each council area, to identify relevant sites.
- We examined the City Council website for smokefree policies and these typically included mention of specific pedestrianised plazas, malls and boulevards (for references to website links see Table 1).
- We conducted a Google search to identify news items around smokefree areas within each city. For example, some documents specifically identified city plazas (eg, for Hastings^37^).
- Sites that were outside of a council’s jurisdiction were excluded (eg, on private land, university and hospital campuses, and part of national monuments etc). We also excluded any of the potential sites if construction was underway at the time of the field visit.

**Table 1:** City Councils (CC) and District Councils (DC) in the sample and how their smokefree policies relate to outdoor pedestrian-only plazas/malls/boulevards

| **Council (north to south)**<br>**(reference to smokefree policy)** | **Year last updated** | **Covers outdoor plazas or malls/boulevards in some way** | **Policy details signage requirements or mentions (for any outdoor smokefree area)** | **Comment on specifics that may related to pedestrian-only plazas/malls/boulevards** |
| --- | --- | --- | --- | --- |
| Napier CC<sup>39</sup> | 2016 | Yes | "Signage promoting positive smokefree messages will be installed in appropriate places." | The smokefree policy includes: "Council owned urban parks, sportsgrounds, playgrounds and reserves, excluding beach reserves"; "Areas set up primarily for café or dining purposes on publicly-owned land; and Council owned tables in public areas." It is unclear if the non-café table part of Market St is considered to be an urban park. However, the smokefree signs on the wooden council seating suggests that the Council intends this area to be smokefree so this is how we treated this site in our analysis. |
| Hastings DC<sup>39</sup> | 2016 | Yes | See above | See above (policy combined with Napier CC one). Also specifically covered is: "Hastings City Square / Central Plaza". |
| Palmerston North CC<sup>40</sup> | 2020 | Yes | "Council will provide Smokefree/ Auahi kore and vapefree signage for all places designated 'smokefree' and 'vapefree' under this policy." | The policy states smokefree/vapefree for: "streets in the city centre" and "parks and playgrounds". |
| Masterton DC<sup>41</sup> | 2017 | Yes | "This policy will be implemented through the placement of smokefree signs at designated non-smoking areas." | The smokefree policy covers: "MDC owned parks", "council owned seating in public areas"; and "areas set up primarily for café or dining purposes in publicly owned land". ... "Purpose of the Smokefree Policy is to reduce the visibility of smoking in the Masterton district and promote a clean, safe, healthy environment for our community." |
| South Wairarapa DC<sup>42</sup> | 2015 | Unclear | Not mentioned | The smokefree policy covers: Council owned buildings, Council swimming pools, coastal reserves and a stadium and "Any other facility considered to be controlled by the Council". The policy is intended to "protect the community and in particular all persons working in or around Council owned or controlled buildings and facilities". The Greytown |
|  |  |  |  | Town Centre is covered, but this is a building, not an outside area. It is unclear if “facilities” controlled by the Council include other outdoor spaces that are not specifically mentioned in the policy. |
| Porirua CC<sup>43</sup> | 2019 | Yes | “Signage will be provided in English and Te Reo reflecting our bi-lingual signage policy. Relevant Managers will review our properties, parks, playgrounds etc and identify what additional signage is needed in new and existing smokefree areas.” | The Council smokefree policy covers “City Centre/Cobham Court” and “Within 10 metres of the entrance to Council owned facilities, including but not limited to [amongst others]: Council administration building; Pātaka Art + Museum/main library; Te Rauparaha Arena/Aquatic Centre Complex.” “For the purpose of this policy e-cigarettes and vaping are treated as tobacco products and as such are banned in smokefree areas” |
| Upper Hutt DC<sup>44</sup> | 2020 | Yes (in city centre) | “The focus for signage is firstly on areas where children and families congregate or socialise, and areas where smoking rates are high. Implementation of the policy includes signage and messaging in Te Reo Māori.” “Signage will include a ‘no vaping’ message where appropriate”; “signage .... [will] include cessation support messaging where appropriate”. | Policy includes: “Within nine metres of outdoor public areas around Council buildings and facilities”, “Outdoor dining and drinking areas on footpaths”, “Outdoor public areas in the city centre”. “Council, through its publicity and communication, asks that people not vape in smokefree spaces or at smokefree events.” |
| Hutt CC<sup>45</sup> | 2019 | Yes | “The focus for signage is firstly on the most popular areas, areas where children and families congregate or socialise, and areas where smoking rates are high. Implementation of the policy includes signage and messaging in Te Reo Māori.” “The Council, through its publicity and communication, asks that people not vape in smokefree spaces or at smokefree events. Signage does not include a “no vaping” message.” | Policy includes: “Parks” “Outdoor pavement dining areas; Suburban centres and the CBD.” “Outdoor public areas around Council buildings and facilities ... Outdoor pavement dining areas”. |

| Council (north to south)<br>(reference to smokefree policy) | Year last updated | Covers outdoor plazas or malls/boulevards in some way | Policy details signage requirements or mentions (for any outdoor smokefree area) | Comment on specifics that may related to pedestrian-only plazas/malls/boulevards |
| --- | --- | --- | --- | --- |
| Wellington CC<sup>46</sup> | 2019 | Yes | “Provide signs in smokefree outdoor spaces where it is practical to do so, and in line with best practice for Council signs and effective smokefree signs.” | “Civic Precinct and Civic Square (including all public entrances) - All public entrance ways out to 10 meters”, “Grey Street pocket park” (ie, the “pedestrian area between Grey Street and Lambton Quay”). Laneways: Includes Eva Street, Leeds Street, Egmont Street and parts of “Chew’s Lane”, “Midland Park”, and “Waitangi Park”. “The Council asks that people not vape in smokefree spaces or at smokefree events.” |
| Queenstown-Lakes DC<sup>47</sup> | 2006 | No | Not mentioned | The smokefree policy was adopted in 2006 and covers Council-owned playgrounds, sports fields and swimming pools. Therefore, none of the identified pedestrian-only plazas/malls/boulevards with permanent seating were officially designated as smokefree. A new policy is being drafted (in 2021) and the Council trialled smokefree and vape-free waterfronts in Queenstown, Frankton, Glenorchy, and Wanaka from 16 December 2019 to 31 March 2020 as part of the development of a new, comprehensive smokefree policy. |

### Data collection at site visits

On field visits to each site we identified the presence or absence of the features required for inclusion (see above). For sites meeting such criteria, we then collected data on:

- The presence of all smokefree and vapefree signage (with photographs taken of all signs). For the study, we did not include data on smokefree signage which was in the corner of warning signs that were predominantly for other purposes such as “no alcohol” or “no skateboarding” signs. The latter applied to two sites, with a small smokefree logo in the corner of such signs. We also did not include smokefree signage on temporary tables at the site.
- The presence or not of dining/drinking tables, including temporary ones associated with cafés, restaurants and pubs.
- The presence or not of ash trays on any of the tables or special bins for cigarette butts.

Three sites were examined by two authors together to confirm the feasibility of the definitions and methods. Then the rest of the sites were examined by each of the authors alone, albeit with site photographs examined by the other authors. All site visits were conducted during business hours to ensure that any cafés/restaurants and pubs would have any tables set up outside. The survey covered the period 2 January 2021 to 13 May 2021. All the the raw data are available on request from the first author.

## RESULTS

Of the 10 councils, all had details of smokefree outdoor policies on their websites (Table 1). There was also evidence of specific commissioned research on the topic that had been published.^38^ The average time since the smokefree policy was last updated was four years (ie, in 2017; range 2006 to 2020; Table 1). Policies with components covering smokefree outdoor plazas/malls/boulevards were common (80%; 8/10 councils), with the policy of one of the remainder being classified as “unclear”. But only five councils (50%) included a vapefree policy for the outdoor areas covered. Most councils (80%) had some policy around smokefree signage. In three of these (30%), the signage policy specifically mentioned signage in te reo Māori, but only two mentioned vapefree signage. The Hutt City policy specifically stated: “Signage does not include a ‘no vaping’ message”.

A total of 60 relevant pedestrianised sites with permanent seating were identified and surveyed (52 plazas, 6 malls, and 2 boulevards) (Table 2). Of these 63% were officially designated smokefree in the council’s smokefree policy. Smokefree signage was only present in 15% (9/60) of the total sites and in 24% (9/38) of the designated smokefree sites. In these designated sites, the average number of smokefree signs was 1.4 (range: 0 to 14; Table 2). In the sites with smokefree signage, 44% had at least some signs with te reo Māori wording (eg, Figure 1) and 22% had some signs where the smokefree message was just part of a larger sign (eg, Figure 2). There were no vapefree signs at any of the sites and no signs mentioned any enforcement details (eg, telephone numbers for complaints). Qualitative issues included some very small smokefree signs (under 10 cm by 10 cm in area; eg, Figure 2), and some signs were located where the intent was unclear (eg, on the side of a building where the sign could potentially be interpreted as indicating a smokefree *indoor* area).

**Table 2:** Results for pedestrianised sites (plazas/malls/boulevards) in the 10 Council areas (with site specific details available on request from the authors)

| Variable | Key result | Numbers | Additional details |
| --- | --- | --- | --- |
| ***Designated smokefree status by site type*** | **% Smokefree** |  |  |
| Plazas | 65.4% | 34/52 |  |
| Malls | 50.0% | 3/6 |  |
| Boulevard | 50.0% | 1/2 |  |
| Total of the above sites | 63.3% | 38/60 | One was only partially smokefree eg, just the area with tables. |
| ***Ash trays*** | **%** |  |  |
| Ash trays when tables present | 12.0% | 3/25 | 5 of these 25 tables were temporary tables. There were no ash trays at designated smokefree sites |
| ***Smokefree signage*** | **% or number** |  |  |
| Any smokefree signs at all the sites | 15.0% | 9/60 |  |
| Any smokefree signs at designated smokefree sites | 23.7% | 9/38 |  |
| Average number of signs in designated smokefree sites | 1.4 signs |  | Median = 0; range = 0 to 14 per site. There was a statistically significant difference in signage presence between designated smokefree sites and non-designated sites (p=0.015, 1-tailed test; Kruskal-Wallis test) |
| ***For sites with any smokefree signs*** | **% or number** |  |  |
| Average number of different types of signs per site | 1.7 signs |  | Median = 1, range = 1 to 4 per site |
| Presence of any signs with any te reo Māori wording | 44.4% | 4/9 | Mean of 2.1 signs in te reo per site; median = 0, range = 0 to 14 per site. These included a sign with just "Aotearoa" as the only word in te reo. See Figure 1 for an example. |
| Presence of any smokefree signs that were part of other signs | 22.2% | 2/9 | See Figure 2 for an example. |
| Presence of any signs also banning vaping | 0.0% | 0/9 |  |
| Presence of any signs that refer to enforcement details (eg, telephone number for complaints) or fines | 0.0% | 0/9 |  |

**Figure 1:**
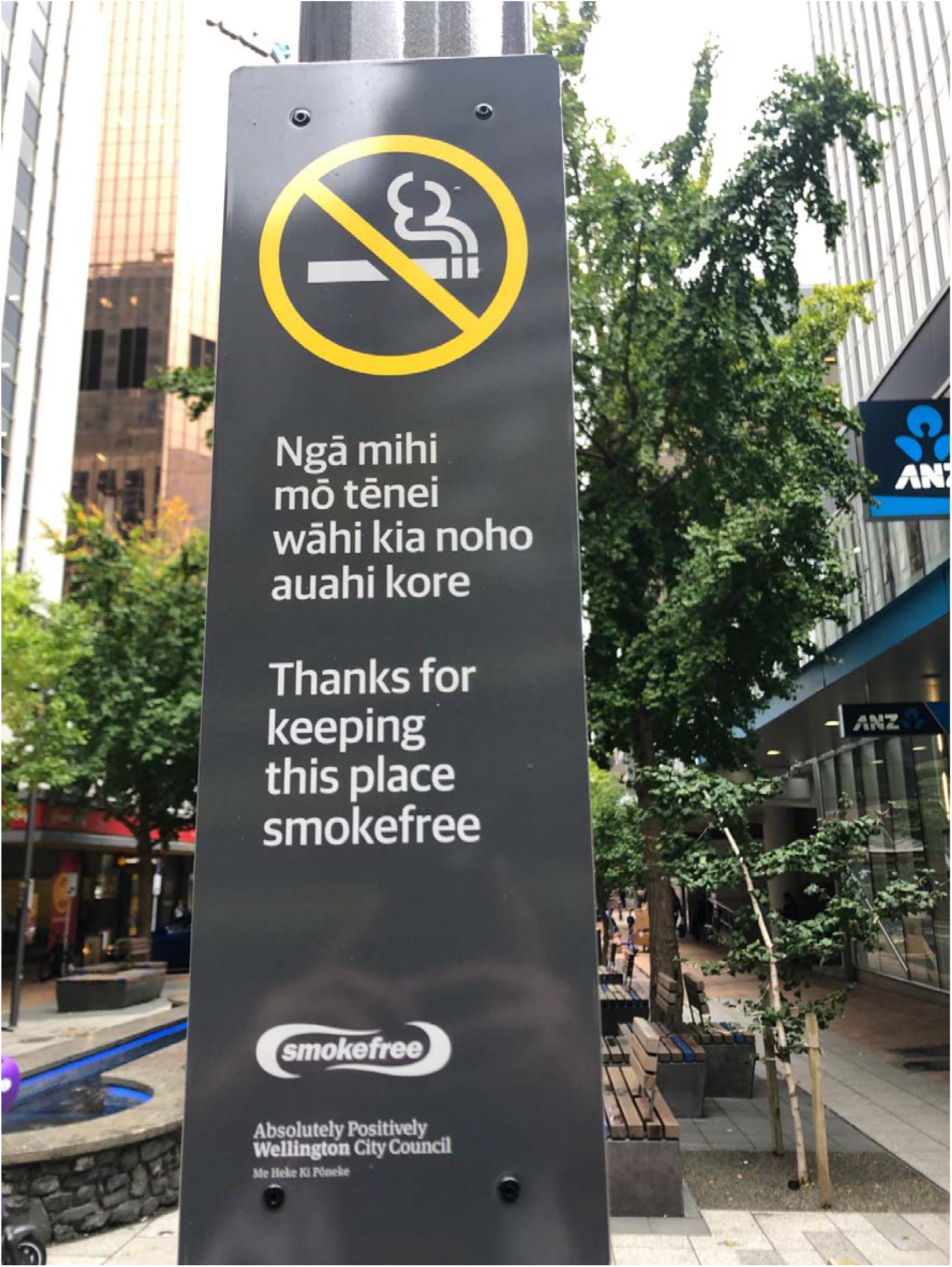
Example of a relatively large smokefree sign with a clear “no smoking” symbol and prominent use of te reo Māori text (photograph by the second author)

**Figure 2:**
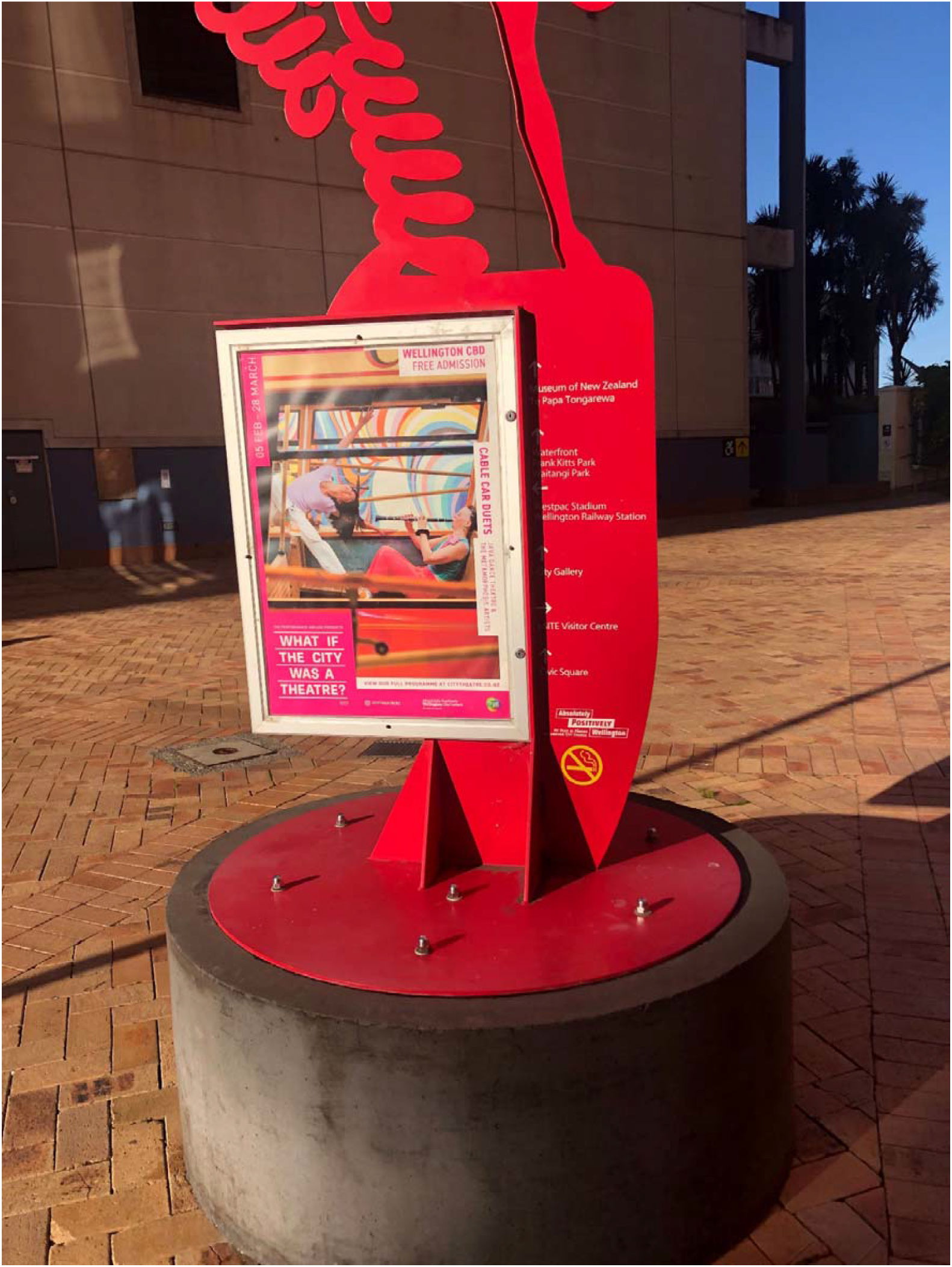
Example of a small smokefree sign being part of a much larger sign for another purpose (the smokefree symbol is in the lower right side of the sign; photograph by the second author)

At sites where tables were present, 12% had ash trays on the tables. But none of the designated smokefree sites had ash trays present on the tables.

## DISCUSSION

A key finding of this survey was that all the 10 councils had details of smokefree outdoor policies on their websites and most (80%) of these policies had components that covered smokefree outdoor plazas/malls/boulevards. Nevertheless, out of the 60 pedestrianised sites with permanent seating that we examined, only 63% were officially designated smokefree (and in some cases this status was unclear or just applied to part of the site). Furthermore, only 24% of the designated smokefree sites had any smokefree signage. This signage was often suboptimal in terms of size, extent, and quality. Also, the 12% of the sites with tables that had ash trays potentially give a problematic pro-smoking signal.

These various problems that we found are inconsistent with the New Zealand Government’s Smokefree Aotearoa 2025 Goal,^32^ as indeed are the extensive limitations with other outdoor smokefree areas in the country (see *Introduction*). They are also inconsistent with local government efforts to make their localities more attractive to workers, shoppers, and tourists; to have a healthier and more productive population;^48^ and to improve the environment with reduced tobacco-related litter and cleaning costs.^49^

Given these problems, there is a case for an upgrade to national smokefree legislation, as is used for indoor public and work places. Such legislation appears to have worked well for another type of outdoor area in New Zealand (ie, school grounds throughout the country^23^), and a national approach has recently been taken with smokefree vehicles.^50^ In contrast to local government initiatives, national legislation has the advantage of providing consistent messaging throughout the country, allowing integration with national smokefree media campaigns, and allowing for economies of scale with centralised sign production/distribution. Specific features of a new national law smokefree law that encompassed these sites could state that:

- All outdoor pedestrianised plazas, malls and boulevards that have any permanent seating are smokefree and vapefree (along with a 10 metre zone from their boundaries).
- All these sites are required to have smokefree signage that meet minimum government specifications (ie, for number of signs per area, size, use of te reo Māori, and messaging including “no vaping”).

Nevertheless, until a national approach is potentially adopted, local government can still take additional initiatives to upgrade their smokefree policies and build them into bylaws. The range of approaches for the use of locally-based laws for smokefree outdoor areas in New Zealand has been detailed previously.^51^

A strength of our study was that it appears to be the first survey (to our knowledge) of the smokefree status these type of outdoor pedestrianised areas in Australasia. Study limitations include the sample of just 10 council areas (out of a potential 67 territorial authorities in the country) and the sample being a convenience one, owing to this being an unfunded study. Also, within the council areas it is possible that we may have missed identifying some of the largest 5-10 sites, owing to our mechanisms for identifying them (eg, via Google Maps, Google Street View and local observations).

In summary, smokefree plazas/malls/boulevards in this survey had multiple policy and signage deficiencies that are inconsistent with achieving the national smokefree goal for 2025. There is scope to address these issues and others highlighted in research on smokefree outdoor policies in Aotearoa, with a major upgrade to the national smokefree law to help denormalise smoking and to help ex-smokers stay quit. This fundamental move on smokefree policies would be in line with the innovative ideas in the Government’s proposals for a Smokefree 2025 Action Plan.^35^

## Data Availability

The raw data are available on request from the first author.

## References

1. Sureda X, Bilal U, Fernandez E, Valiente R, Escobar FJ, Navas-Acien A, Franco M. Second-hand smoke exposure in outdoor hospitality venues: Smoking visibility and assessment of airborne markers. Environ Res 2018;165:220–27.

2. Pearson AL, Nutsford D, Thomson G. Measuring visual exposure to smoking behaviours: a viewshed analysis of smoking at outdoor bars and cafes across a capital city’s downtown area. BMC Public Health 2014;14:300.

3. Zablocki RW, Edland SD, Myers MG, Strong DR, Hofstetter CR, Al-Delaimy WK. Smoking ban policies and their influence on smoking behaviors among current California smokers: a population-based study. Prev Med 2014;59:73–8.

4. Chaiton M, Diemert L, Zhang B, Kennedy RD, Cohen JE, Bondy SJ, Ferrence R. Exposure to smoking on patios and quitting: a population representative longitudinal cohort study. Tob Control 2016;25:83–8.

5. Johns M, Coady MH, Chan CA, Farley SM, Kansagra SM. Evaluating New York City’s smoke-free parks and beaches law: a critical multiplist approach to assessing behavioral impact. Am J Community Psychol 2013;51:254–63.

6. Johns M, Farley SM, Rajulu DT, Kansagra SM, Juster HR. Smoke-free parks and beaches: an interrupted time-series study of behavioural impact in New York City. Tob Control 2014,

7. Breunis LJ, Bebek M, Dereci N, de Kroon MLA, Rado MK, Been JV. Impact of an inner-city smoke-free zone on outdoor smoking patterns: a before-after study. Nicotine Tob Res 2021,

8. Thomson G, Wilson N, Collins D, Edwards R. Attitudes to smoke-free outdoor regulations in the USA and Canada: a review of 89 surveys. Tob Control 2016;25:506–16.

9. Sureda X, Fernandez E, Martinez-Sanchez JM, Fu M, Lopez MJ, Martinez C, Salto E. Secondhand smoke in outdoor settings: smokers’ consumption, non-smokers’ perceptions, and attitudes towards smoke-free legislation in Spain. BMJ Open 2015;5:e007554.

10. Walsh R, Paul C, Tzelepis F, Stojanovski E, Tang A. Is government action out-of-step with public opinion on tobacco control? Results of a New South Wales population survey. Aust N Z J Public Health 2008;32:482–8.

11. Rosenberg M, Pettigrew S, Wood L, Ferguson R, Houghton S. Public support for tobacco control policy extensions in Western Australia: a cross-sectional study. BMJ Open 2012;2:e000784.

12. Brenner G, Mann K, Lee D, Burrows J, Crosby S. Attitudes towards smokefree high streets: a survey of local shoppers in a northern UK town. Perspect Public Health 2018;138:325–28.

13. Wilson N, Blakely T, Edwards R, Weerasekera D, Thomson G. Support by New Zealand smokers for new types of smokefree areas: national survey data. N Z Med J 2009;122(1303):80–9.

14. Gallus S, Rosato V, Zuccaro P, Pacifici R, Colombo P, Manzari M, La Vecchia C. Attitudes towards the extension of smoking restrictions to selected outdoor areas in Italy. Tob Control 2012;21:59–62.

15. Martinez C, Guydish J, Robinson G, Martinez-Sanchez JM, Fernandez E. Assessment of the smoke-free outdoor regulation in the WHO European Region. Prev Med 2014;64:37–40.

16. American Non-Smokers Rights Foundation. Municipalities with Smokefree Outdoor Dining and Bar Patio Laws. Berkeley: American Nonsmokers’ Rights Foundation, 2021,

17. Center for Tobacco Policy & Organizing. Comprehensive outdoor secondhand smoke ordinances. Sacramento: Center for Tobacco Policy & Organizing: American Lung Association of California, 2012,

18. Thomson G. Smokefree Wellington: Context, options and evidence. Wellington: University of Otago, Wellington, 2015,

19. New York City Department of Parks & Recreation. Smoke-Free Parks and Beaches. New York: New York City Department of Parks & Recreation, 2021,

20. Institute of Health Metrics and Evaluation. GHDx (Global Health Data Exchange); Results for New Zealand (on 24 May 2021,. Seattle; Institute of Health Metrics and Evaluation. http://ghdx.healthdata.org/gbd-results-tool.

21. Marsh L, Robertson LA, Kimber H, Witt M. Smokefree outdoor areas in New Zealand: how far have we come? N Z Med J 2014;127(1389):51–66.

22. Thomson G, Wilson N. Local and regional smokefree and tobacco-free action in New Zealand: highlights and directions. N Z Med J 2017;130:89–101.

23. Wilson N, Oliver J, Thomson G. Ten years of a national law covering smoke-free school grounds: a brief review. Tob Control 2016;25:122.

24. Wilson N, Delany L, Thomson GW. Smokefree laws and hospitality settings: an example from New Zealand of a deficient approach. Tob Control 2020;29:460.

25. Wilson N, Thomson G, Edwards R. The potential of Google Street View for studying smokefree signage. Aust N Z J Public Health 2015;39:295–6.

26. Thomson G, Wilson N. Smokefree signage at children’s playgrounds: Field observations and comparison with Google Street View. Tob Induc Dis 2017;15:37.

27. Wilson N, Thomson G. Suboptimal smokefree signage at some hospitals: Field observations and the use of Google Street View. N Z Med J 2015;128(1415):56–9.

28. Thomson G, Wilson N. Smokefree signage at New Zealand racecourses and sports facilities with outdoor stands. N Z Med J 2017;130:80–86.

29. Wilson N, Thomson G. Smokefree signage at railway stations: a survey of 54 stations in 11 local government areas. N Z Med J 2019;132:59–61.

30. Wilson N, Jones AC, Thomson GW. Poor smoke-free status of airports in a country with a smoke-free goal: New Zealand. Tob Control 2020,

31. Wilson N, Thomson G. Surveying all outdoor smokefree signage in contrasting suburbs: methods and results. Health Promot J Austr 2017;28:264–65.

32. New Zealand Government. Government Response to the Report of the Māori Affairs Committee on its Inquiry into the tobacco industry in Aotearoa and the consequences of tobacco use for Māori (Final Response). Wellington: New Zealand (NZ) Parliament, 2011, http://www.parliament.nz/en-nz/pb/presented/papers/49DBHOH_PAP21175_1/government-final-response-to-report-of-the-m%c4%81ori-affairs

33. Gurram N, Thomson G. The point prevalence of smoking and vaping in downtown locations in Wellington: Report for the Wellington City Council on observations in November 2018, Wellington: University of Otago Wellington, November 29, 2018. https://www.otago.ac.nz/wellington/departments/otago711611.pdf

34. Thomson G, Pathamanathan N. The point prevalence of smoking in selected sports fields and downtown locations in Wellington: Observations in November 2015. University of Otago, Wellington, January 2016, http://www.otago.ac.nz/wellington/otago610430.pdf

35. Ministry of Health. Proposals for a Smokefree Aotearoa 2025 Action Plan: Discussion document. Wellington: Ministry of Health 2021, https://www.health.govt.nz/system/files/documents/publications/proposals_for_a_smokefree_aotearoa_2025_action_plan-final.pdf

36. Daube M, Maddox R. Impossible until implemented: New Zealand shows the way. Tob Control 2021,

37. Hastings District Council. Hastings City Centre Revitalisation Plan. Hastings District Council, 2019(February). https://www.hastingsdc.govt.nz/assets/Document-Library/Plans/Hastings-City-Centre-Public-Spaces-Revitalisation-Plan/Hastings-City-Centre-Public-Spaces-Revitalisation-Plan.pdf.

38. Gurram N. The point prevalence of smoking and vaping in Grey Street pocket square, Wellington: Report for the Wellington City Council on observations in Grey Street, Wellington in December 2018. 2019, https://www.otago.ac.nz/wellington/departments/otago711699.pdf.

39. Hastings District and Napier City Councils. Hastings District and Napier City Councils Smokefree Policy Napier: Hastings District and Napier City Councils, 2016,

40. Palmerston North City Council. Smokefree Outdoor Areas Policy. Palmerston North, NZ: Palmerston North City Council, 2020,

41. Masterton District Council. Smokefree policy. Masterton: Masterton Disctrict Council, 2017, https://mstn.govt.nz/wp-content/uploads/2017/02/Smokefree-Policy_Current.pdf.

42. South Wairarapa District Council. Smoke-Free Environment Policy. Martinborough: South Wairarapa District Council, 2015, https://swdc.govt.nz/wp-admin/admin-ajax.php?action=wpmf_onedrive_business_download&id=01CTNIGYECBXJBHTTMBBB3QHHUKFT6IOL7&link=true&dl=0.

43. Porirua City Council. Smokefree outdoor places policy. Porirua: Porirua City Council, 2019,

44. Upper Hutt City Council. Smokefree Policy. (Accessed 5 April 2021). https://www.upperhuttcity.com/Your-Council/Plans-policies-bylaws-and-reports/Policies-and-bylaws/Smokefree-Upper-Hutt.

45. Hutt City. Smokefree Lower Hutt. (Updated 20 July, 2020). http://www.huttcity.govt.nz/Our-City/live-here/smokefree-lower-hutt/.

46. Wellington City Council. Smokefree Wellington. (Accessed 17 January, 2021). https://wellington.govt.nz/your-council/plans-policies-and-bylaws/policies/smokefree-wellington.

47. Queenstown Lakes District Council. Smokefree and Vapefree. 2021,(accessed 12 April) Available from: https://letstalk.qldc.govt.nz/smokefree.

48. Hahn EJ. Smokefree legislation: a review of health and economic outcomes research. Am J Prev Med 2010;39:S66–76.

49. Torkashvand J, Farzadkia M. A systematic review on cigarette butt management as a hazardous waste and prevalent litter: control and recycling. Environ Sci Pollut Res Int 2019;26:11618–30.

50. Wilson N, Thomson G, Edwards R. Smoke-free cars legislation: it works but New Zealand should still rigorously evaluate its upcoming law. N Z Med J 2020;133:99–103.

51. Thomson G, Martin J, Gifford H, Parata K. Expanding smokefree outdoor areas in Wellington City: Rationale and options. Wellington: University of Otago, 2016,

